# Automated Assessment of OSCE Physical Exams using Multimodal AI

**DOI:** 10.64898/2026.01.09.26343786

**Authors:** Shinyoung Kang, Michael J. Holcomb, Ameer H. Shakur, David Hein, Huong-Tra Ngo, Hunter Schuler, Philip C. Jarrett, Thomas O. Dalton, Andrew R. Jamieson

**Affiliations:** Lyda Hill Department of Bioinformatics, University of Texas Southwestern Medical Center. Dallas; Emergency Medicine, University of Texas Southwestern Medical Center. Dallas; Department of Internal Medicine, University of Texas Southwestern Medical Center. Dallas

## Abstract

**Background:** The assessment of physical examination skills in medical education is resource-intensive and prone to inter-rater variability. While artificial intelligence (AI) has successfully automated the grading of clinical notes and transcripts, evaluating the physical techniques themselves—what students do rather than what they say—remains an unsolved challenge. We evaluated whether a multimodal AI system could assess physical examination skills with expert-level reliability.

**Methods:** In this retrospective ablation study, we analyzed 300 video-recorded encounters from six Objective Structured Clinical Examination (OSCE) stations (cardiovascular, respiratory, gastrointestinal, musculoskeletal, and neurological). We compared the performance of a multimodal AI model (Gemini 2.5 Pro) across single- and multi-camera configurations and isolated input modalities (video, audio-only, transcript-only, visual-only) against standard human grading. The primary outcome was agreement with a physician-adjudicated ground-truth reference standard, measured by quadratic weighted Cohen’s kappa (κ).

**Results:** The AI system using a synchronized 3-camera native video configuration achieved significantly higher reliability (κ = 0.830; 95% CI, 0.773–0.880) than the standard human evaluators (κ = 0.732; 95% CI, 0.687–0.776). Performance followed a strict hierarchy: native video > audio-only > transcript-only > visual-only. Notably, visual-only models failed (κ ≈ 0.20) despite high detection accuracy, revealing a “visual paradox” where models could identify when an action occurred but not how well it was performed without audio cues.

**Conclusions:** A properly configured multimodal AI system can grade physical examination skills with reliability exceeding that of trained human evaluators. Success requires native processing of synchronized audio-visual streams; transcript-based or visual-only approaches are insufficient for high-stakes assessment. These findings suggest that AI can provide scalable, objective, and valid assessment of clinical skills, overcoming the limitations of traditional human grading.

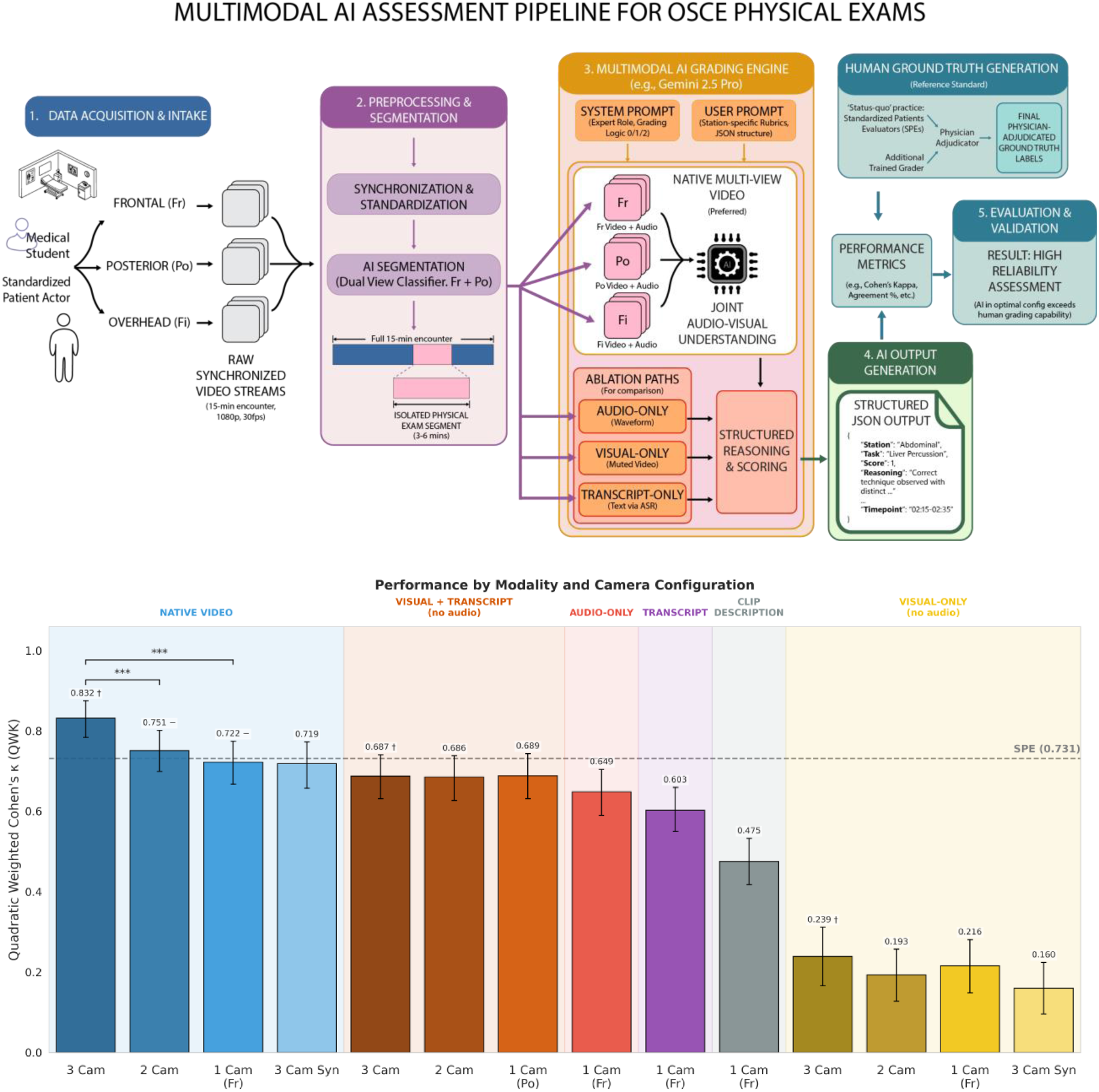

## Introduction

The Objective Structured Clinical Examination (OSCE) has emerged as the gold standard for assessing clinical competencies in medical education, providing a standardized approach for evaluating students’ practical skills across diverse clinical scenarios [1]. During medical school, this examination allows students to demonstrate their abilities through immersive, real-time patient encounters with trained actors known as standardized patients (SPs). These scripted encounters provide students with realistic experiences while maintaining the consistency needed for fair assessment. Typically, an OSCE consists of multiple 15- to 20-minute-long encounters or “stations”. Audio-visual recording of these encounters is a common practice to support student evaluations. The encounter recordings provide a rich data stream for evaluating student performance, capturing communication, body language, emotional affect, and physical examination actions [2].

However, the traditional OSCE assessment process is resource-intensive, requiring both specific medical expertise and sustained attention to detail for extended periods [3]. These constraints impose practical limitations on what can be reasonably implemented, incentivizing simplified and often underspecified rubrics. When exams are conducted across multiple stations and entire student cohorts, challenges with evaluator fatigue and inter-rater reliability intensify [4, 5]. Furthermore, medical schools often rely on trained, non-physician staff to conduct grading [6]. Hence, despite the time and resources committed to conducting these elaborate, data-rich exercises, the task of returning high-quality assessments to students in a timely fashion remains daunting [3–6].

Audio-visual recordings of OSCE may enable AI evaluation of student performance to alleviate human workload. Recent advances in AI-driven OSCE assessment have shown promise, particularly for evaluating clinical documentation and communication skills. Large language models (LLMs) have been successfully deployed to grade OSCE post-encounter notes, achieving up to 89.7% agreement with human expert graders at the rubric item level and dramatically reducing grading turnaround time from weeks to days [7]. Similarly, OSCE transcripts have been analyzed with LLMs to assess communication and clinical reasoning, demonstrating encouraging initial results [8, 9].

Yet these successes have been limited to verbal and written components of clinical examinations, leaving a critical gap in the assessment of physical examination skills, a core component of clinical competence that requires visual observation and evaluation of procedural techniques. While audio-based data streams fed to AI can directly address clinical competence related to spoken language and verbalized communication, visual observation is required for proper assessment of physical examination skills and procedural techniques.

The assessment of physical examination skills presents unique challenges that have thus far proven resistant to AI automation [10, 11] .Unlike skills that can be captured through transcripts or clinical reasoning that manifests in verbal explanations, physical examination techniques require precise observation of hand placement, palpation technique, examination sequence, and patient interaction dynamics. Initial attempts to automate physical examination assessment have shown limited success [10]. Transformer models have been used for context-aware video segmentation to detect whether specific physical examinations occurred during OSCE encounters, representing an essential first step toward automation. However, this work focused solely on binary detection of physical examination content rather than quality assessment.

Attempts to grade the quality of physical examinations using multimodal question-answering models found that transcript-only models paradoxically outperformed multimodal approaches incorporating visual inputs [11]. The finding that incorporating visual information degrades performance suggests that multimodal AI systems struggled to accurately interpret physical examination techniques, instead relying heavily on conversational cues from student – patient dialogue to interpret student actions while generating inaccurate visual interpretations that confused the assessment process. Notably, these approaches did not employ preprocessing to isolate examination segments, potentially allowing irrelevant video content to interfere with assessment accuracy.

This progression from detection [10] to limited grading attempts [11] highlights a fundamental challenge in medical education technology: while AI has proven capable of assessing what students say or write, and can detect whether physical examinations occur, it has not yet reliably evaluated the quality of what students do during physical examinations. This limitation is particularly problematic given that physical examination skills represent a critical component of clinical competence that cannot be adequately assessed through verbal proxies alone. The disconnect between verbal signposting and actual examination technique quality creates a significant validity threat, as students who excel at describing procedures may not necessarily perform them correctly.

This study addresses these challenges through a systematic ablation analysis of multimodal AI for automated physical examination assessment in OSCEs (Figure 1). Our objectives are threefold:

**Figure 1.**
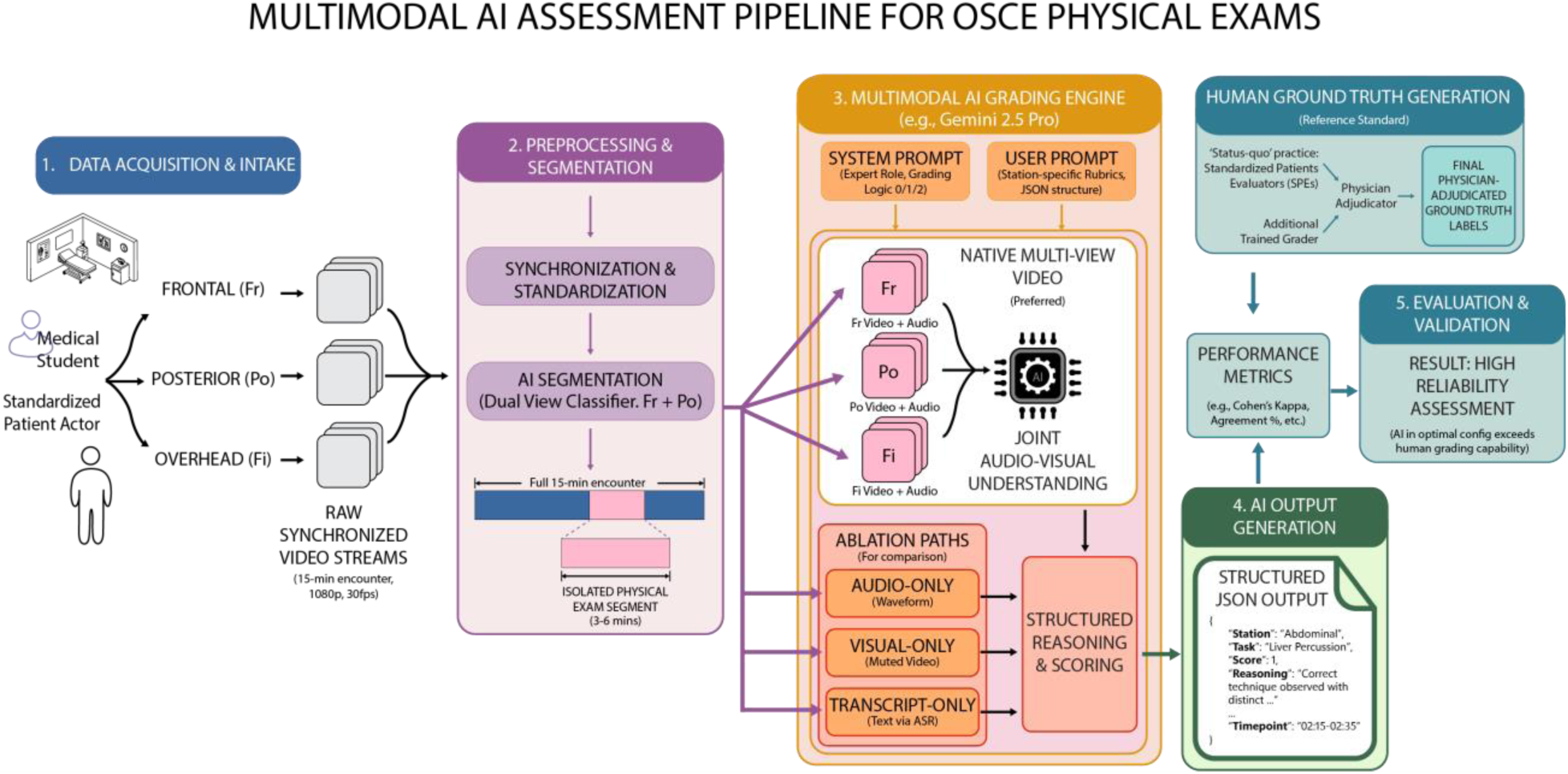
Schematic for Multimodal AI Assessment Pipeline for OSCE Physical Exams.

First, we evaluate whether AI can achieve expert-level agreement in grading physical examination skills by comparing automated assessments against expert human evaluators across 300 OSCE examinations from six different clinical stations.

Second, we determine the relative contribution of different input modalities—video (combined audio-visual), audio-only, visual-only, transcript-only, and combinations thereof—to establish which modalities are necessary and sufficient for accurate physical examination assessment.

Third, we quantify the impact of multi-camera configurations by comparing assessment performance using one, two, or three camera angles, providing evidence-based guidance for optimal recording infrastructure.

Through this comprehensive ablation approach, we aim to determine whether properly configured multimodal AI systems can successfully grade physical examination skills, and if so, identify the optimal configuration that balances assessment accuracy with practical implementation considerations.

## Methods

### Physical Examination Assessment

In this retrospective ablation study, we evaluated students’ physical examination performance across six distinct OSCE stations, each designed to assess competencies in different clinical domains. The physical examination component of each station focused on specific procedural skills relevant to the clinical scenario, including cardiovascular, respiratory, neurological, musculoskeletal, and abdominal examinations.

Each station employed standardized rubrics containing around 5 discrete physical examination items tailored to the clinical presentation. Items were selected based on clinical relevance, educational objectives, and the ability to be reliably observed through video recording. The rubrics evaluated both the occurrence and quality of examination maneuvers, with detailed criteria for each assessment level (see Supplemental Material S1 for example rubrics).

Students were assessed using a three-level scoring system for each physical examination item:

- Score 0 (Not Performed): The examination maneuver was not attempted or could not be identified
- Score 1 (Partially Performed): The examination technique was attempted but executed with significant deficiencies in technique, sequence, or completeness
- Score 2 (Correctly Performed): The examination was completed with appropriate technique, proper sequence, and adequate thoroughness

Assessment criteria included proper hand placement, appropriate palpation pressure, correct examination sequence, patient positioning, use of appropriate equipment when indicated, and maintenance of patient comfort and dignity throughout the examination. Verbal signposting of examination steps was noted but not sufficient for credit without corresponding physical performance.

Timing and Documentation: Physical examination segments typically ranged from 3-6 minutes within the 15-minute encounter, with variation based on station complexity and clinical requirements. All physical examination maneuvers were required to be completed within the designated time window. Students received no credit for examinations described but not performed or for maneuvers attempted after time expiration.

### Reference Standard (Ground-Truth Labels)

The items for each OSCE encounter were graded by at least 2 standardized-patient evaluators (SPE) (Table S1). When the SPEs could not reach a majority agreement, an evaluator re-reviewed the specific item and issued an adjudication. This evaluator could be one of the original raters or a different rater. In either case, this evaluator’s adjudication served as the final SPE consensus for that item, called “SPE (Final)”. This score represents the ‘status quo’ score that is traditionally used.

Independently, an emergency medical technician (EMT) also graded the items. Disagreements between SPE (Final) and EMT grades were adjudicated by a physician. This adjudicated score serves as the reference standard (ground-truth label) for all model performance evaluations. This adjudicated score is denoted as “ground truth”.

Ablation Study Configuration Inter-rater agreement among the SPEs, and between SPEs and the Ground Truth was quantified with percent agreement and quadratic weighted Cohen’s κ (QWK) with bootstrap confidence intervals [11–13] (Table S1, S2).

### Dataset

The corpus included 300 encounters from the 2024 OSCE cycle across six clinical stations conducted at UT Southwestern Medical Center’s Simulation Center (Table 1).

**Table 1.** Video count and physical examination duration statistics by station.

| Station | Number of Video | Number of Physical Exam Question | Mean Physical Exam Duration (s, ± std) | Score Distribution (%) | | |
| --- | --- | --- | --- | --- | --- | --- |
|  |  |  |  | 0 | 1 | 2 |
| Overall | 300 | 23 | 207 ± 135 | 9.7 | 14.3 | 76.0 |
| Abdominal Pain | 50 | 4 | 167 ± 49 | 4.0 | 15.5 | 80.5 |
| Back Pain | 50 | 4 | 328 ± 162 | 5.0 | 33.5 | 61.5 |
| Chest Pain | 50 | 3 | 127 ± 43 | 6.0 | 3.3 | 90.7 |
| Falling | 50 | 4 | 153 ± 104 | 3.5 | 4.5 | 92.0 |
| Joint Pain | 50 | 4 | 170 ± 170 | 31.5 | 9.0 | 59.5 |
| Lung - BP | 50 | 4 | 276 ± 74 | 7.5 | 75.5 | 75.5 |

From a cohort of 200 pre-clerkship medical students who completed the examination, we randomly selected 50 students and extracted their videos across all six clinical stations, yielding a total dataset of 300 encounters. Each encounter lasted approximately 15 minutes and contained synchronized recordings from frontal (Fr), posterior (Po), and overhead fish-eye (Fi) camera views. The unit of analysis was the rubric item for each encounter. Data for this study was acquired with Institutional Review Board approval and processed in a FERPA-compliant compute environment.

### Data Preparation

All videos were standardized to 1920×1080 resolution and 30 frames per second (fps), then temporally aligned across the Fr, Po, and Fi views using shared timestamps. Synchronized multi-view combinations of input streams were also created. From the segmented physical-exam interval, we prepared modality-specific inputs as follows:

Gemini 2.5 Pro received synchronized MP4 streams with the native AAC audio tracks. For audio-only or transcript analysis generation, the AAC track was extracted and re-encoded to MP3 with FFmpeg using variable bitrate (VBR) quality 2 (yielding ∼90 kbps on average), preserving the original 48 kHz sampling rate and channel configuration (mono).

For Qwen-3-Omni, a single camera visual input was uniformly sampled at 1 fps and audio was converted to PCM16E at 16 kHz. Data were formatted according to each provider’s specified input requirements, enabling standardized comparisons across models using their preferred file configurations [14, 15].

#### Physical Examination Segmentation Preprocessing

The physical examination interval is segmented out from the full-length encounter video using a dual-view classifier, an extension of prior single-camera approaches [10]. The synchronized Fr and Po views were used. The full-length encounter video was partitioned into non-overlapping 10-second dual-view windows (i.e., 15-minute recording was partitioned into 90 non-overlapping 10-second dual-view windows (Fr and Po)). The dual-view classifier labeled each window as either “exam” or “non-exam”. Then, the physical exam segment was defined as the continuous interval spanning the first through the last window labeled “exam” and was extracted as an uninterrupted Fr/Po segment (i.e., we did not concatenate disjoint clips). Manual verification was performed for 25% of the segmentation boundaries to verify that the full physical examination segment was captured for every encounter [9], achieving a segmentation recall of 99.8%.

### Multimodal Pipeline Design

Our primary approach used Gemini 2.5 Pro (version 06-05) to process segmented videos directly, leveraging native joint audio-visual understanding. We varied two experimental factors: (i) modality—video (synchronized audio+visual), audio-only (native audio understanding without transcription), visual-only (muted video), transcript-only (transcript was derived from the same segmented exam using Gemini 2.5 Flash, version 06-17 2025 (stable)), and visual+transcript (muted video plus the transcript as text); and (ii) camera configuration —single view 1-camera (Fr, Po, Fi), dual view 2-camera (Fr+Po), and triple view 3-camera (Fr+Po+Fi). Because spoken content is identical across cameras, we report audio-only results for a representative source in the main text (camera Fr) and confirm consistency across sources in sensitivity analyses (Table S3).

#### Prompt Construction

System and user prompts were designed a priori by adapting materials currently used to educate SPEs on student OSCE evaluation. No rounds of prompt refinement were conducted during experimentation. The system prompts were consistent across all stations for a given input modality (native video, audio+transcript, etc.). User prompts substituted only the station name and relevant physical examination rubric items. For each station, the model was asked to score all rubric items in a single call using structured outputs. See prompt examples in Supplemental Material S2, S3.

#### Ablation Study Design

We conducted a 6 × 5 factorial study that crossed six data-modality conditions with five camera configurations. The six modality conditions included native video (synchronized audio + visual), audio-only, visual-only (muted video), transcript-only (text derived from audio), visual + transcript (muted video with transcript text), and a synthesis variant that first generated per-stream judgments and then fused them using a large language model. The five camera configurations consisted of frontal (Fr), posterior (Po), overhead (Fi), dual-view (Fr + Po), and tri-view (Fr + Po + Fi) arrangements.

For the multi-camera setups, two integration strategies were evaluated: a native multiview approach in which synchronized streams were processed jointly at inference, and a post-hoc synthesis approach in which individual views were analyzed separately and subsequently merged into a single grade. The native tri-camera configuration served as the reference condition for all planned comparisons. See Table 2 for the complete experimental design and corresponding model configurations.

**Table 2.** Ablation Study Configuration.

| **Table 2. Ablation Study Configuration** |  |  |  |  |  |
| --- | --- | --- | --- | --- | --- |
| **Processing Type** | **Input Camera Configurations** |  |  |  |  |
|  | Frontal (Fr) | Posterior (Po) | Fish-eye (Fi) | Fr + Po | Fr + Po + Fi |
| **Native** |  |  |  |  |  |
| Video | ✓ | ✓ | ✓ | ✓ | ✓ |
| Audio | ✓ | ✓ | ✓ | ✓ | ✓ |
| Visual<br>(Muted Video) | ✓ | ✓ | ✓ | ✓ | ✓ |
| Visual + Transcript | ✓ | ✓ | ✓ | ✓ | ✓ |
| **Transcript** |  |  |  |  |  |
| Full Video | ✓ |  |  |  |  |
| Physical Exam | ✓ |  |  |  |  |
| **Synthesis** |  |  |  |  |  |
| Video |  |  |  | ✓ | ✓ |
| Audio |  |  |  | ✓ | ✓ |
| Visual |  |  |  | ✓ | ✓ |
| **Clip Description** | ✓ |  |  |  |  |

**Table 3.**
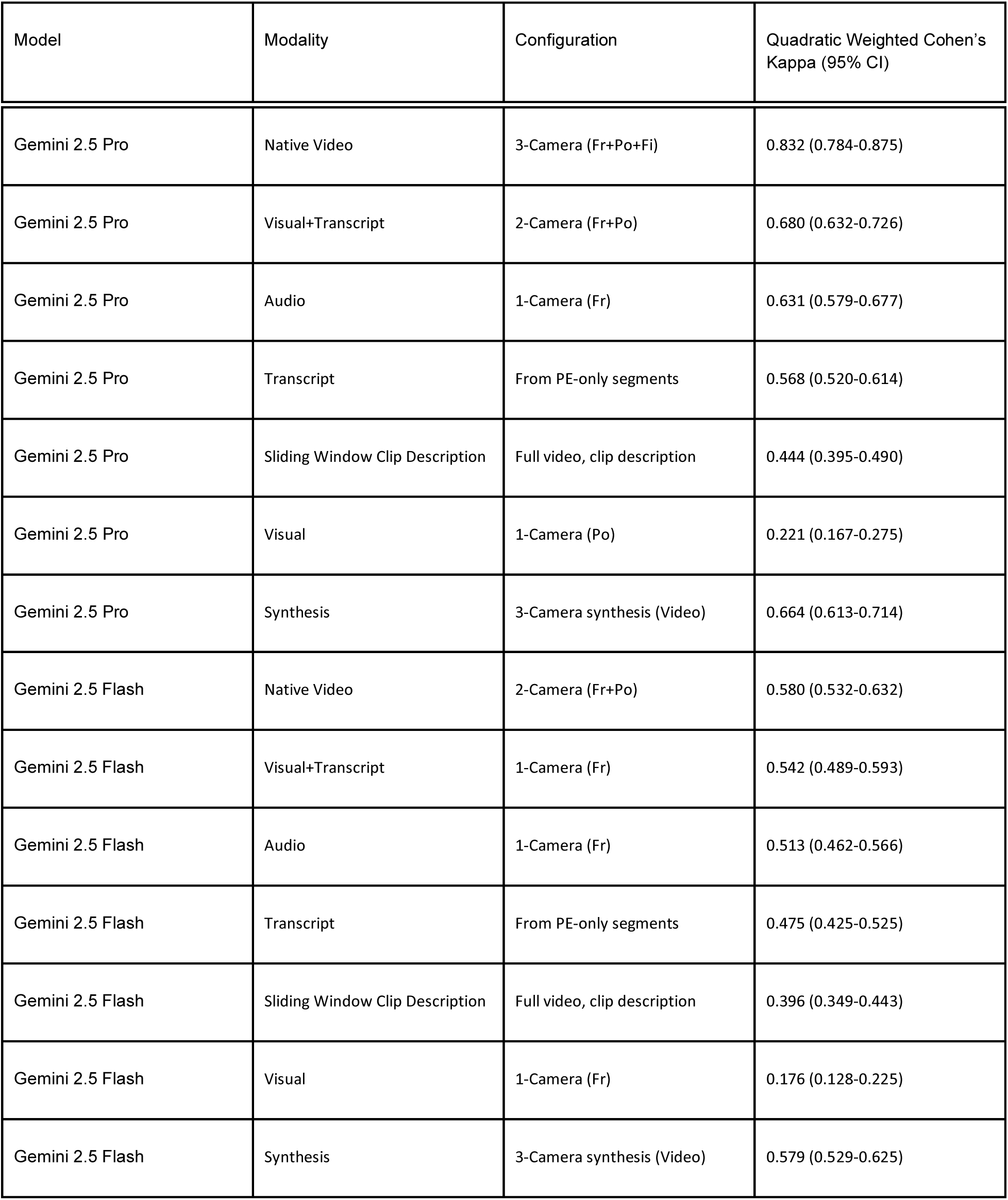
Top Configurations Categorical kappa of each modality.

| Table 3. Top Configurations Categorical kappa of each modality |  |  |  |
| --- | --- | --- | --- |
| Model | Modality | Configuration | Quadratic Weighted Cohen's Kappa (95% CI) |
| Gemini 2.5 Pro | Native Video | 3-Camera (Fr+Po+Fi) | 0.832 (0.784-0.875) |
| Gemini 2.5 Pro | Visual+Transcript | 2-Camera (Fr+Po) | 0.680 (0.632-0.726) |
| Gemini 2.5 Pro | Audio | 1-Camera (Fr) | 0.631 (0.579-0.677) |
| Gemini 2.5 Pro | Transcript | From PE-only segments | 0.568 (0.520-0.614) |
| Gemini 2.5 Pro | Sliding Window Clip Description | Full video, clip description | 0.444 (0.395-0.490) |
| Gemini 2.5 Pro | Visual | 1-Camera (Po) | 0.221 (0.167-0.275) |
| Gemini 2.5 Pro | Synthesis | 3-Camera synthesis (Video) | 0.664 (0.613-0.714) |
| Gemini 2.5 Flash | Native Video | 2-Camera (Fr+Po) | 0.580 (0.532-0.632) |
| Gemini 2.5 Flash | Visual+Transcript | 1-Camera (Fr) | 0.542 (0.489-0.593) |
| Gemini 2.5 Flash | Audio | 1-Camera (Fr) | 0.513 (0.462-0.566) |
| Gemini 2.5 Flash | Transcript | From PE-only segments | 0.475 (0.425-0.525) |
| Gemini 2.5 Flash | Sliding Window Clip Description | Full video, clip description | 0.396 (0.349-0.443) |
| Gemini 2.5 Flash | Visual | 1-Camera (Fr) | 0.176 (0.128-0.225) |
| Gemini 2.5 Flash | Synthesis | 3-Camera synthesis (Video) | 0.579 (0.529-0.625) |

#### Sliding Window Clip Description

For comparison with contemporary “native multimodal” pipelines, we implemented an enhanced version of the Holcomb et al. “storytelling” method [11]. From the segmented videos of the physical examinations, we extracted frames at 1 fps and partitioned the exam into 15-second clips (e.g., a 90-second exam is partitioned into 6 clips of 15 frames and transcript). Based on the 15-frames in each clip, Gemini 2.5 Pro produced structured, clip-level descriptions focused on hand placement, technique, and equipment use. These clip descriptions were interleaved chronologically with Gemini-derived transcripts to form “exam stories,” which were then graded deterministically (temperature = 0). Unlike the earlier work [11] which suffered from an overreliance on correct keyframe identification, this proposed update uniformly samples descriptions from the physical exam, sidestepping this critical dependency on precise action timestamps. (A GPT-4.1 + Whisper v3 variant is reported in the Supplement as a sensitivity check.)

#### Targeted Visual Re-analysis

To isolate whether visual-only model failures stem from temporal localization versus visual interpretation deficits, we conducted a targeted analysis. We identified rubric items where the native tri-camera model assigned a non-zero score (N = 1,034, 90%), indicating that an action was detected. For each item, we extracted the start and end timestamps from the tri-camera model’s output and trimmed the muted video to show only the relevant time window. We compared performance between the standard visual-only condition (full segmented exam video) and the targeted condition (item-specific trimmed video segments) using McNemar’s test for accuracy differences and Wilcoxon signed-rank test for κ differences.

#### Evaluation Framework

Primary outcomes included agreement with the ground truth and quadratic-weighted Cohen’s κ (QWK). QWK was selected as the primary metric because it corrects for chance agreement and explicitly accounts for the ordinal structure of the three-level scoring rubric (0, 1, 2), making it well suited for evaluating grading reliability in OSCE assessments.

Secondary outcomes included macro-averaged F1 and question-level accuracy. Macro-F1 was used as a class-balanced performance measure to facilitate comparison across rubric items with substantial class imbalance, particularly where minority outcomes (e.g., partially performed maneuvers) are educationally meaningful. Because macro-F1 does not account for ordinal severity or chance agreement, it was used for descriptive, item-level analysis rather than for assessing overall grading reliability, making it complementary to primary QWK measure.

### Statistical Analysis

Analyses were performed at the rubric-item level using the physician-adjudicated Ground Truth as the reference standard. Agreement between AI predictions, SPEs, and the Ground Truth was assessed using percent agreement and quadratic weighted Cohen’s κ, the primary measure of chance-corrected reliability across the three-level rubric (0, 1, 2). Bootstrapped 95% confidence intervals (CIs) were generated from 1,000 resamples using student-level clustering Pairwise κ differences across modality and camera configurations were compared using paired resampling tests with Benjamini-Hochberg FDR correction (20 comparisons, α = 0.05).

## Results

Across 300 OSCE examinations, the best-performing AI configuration (Gemini 2.5 Pro 3-camera native video) achieved a QWK Cohen’s κ = 0.832 (95% CI: 0.784 - 0.875) agreement with Ground Truth compared with human grader (SPEs) performance at κ = 0.731 (95% CI: 0.692 - 0.764) (Figure 2).

**Figure 2.**
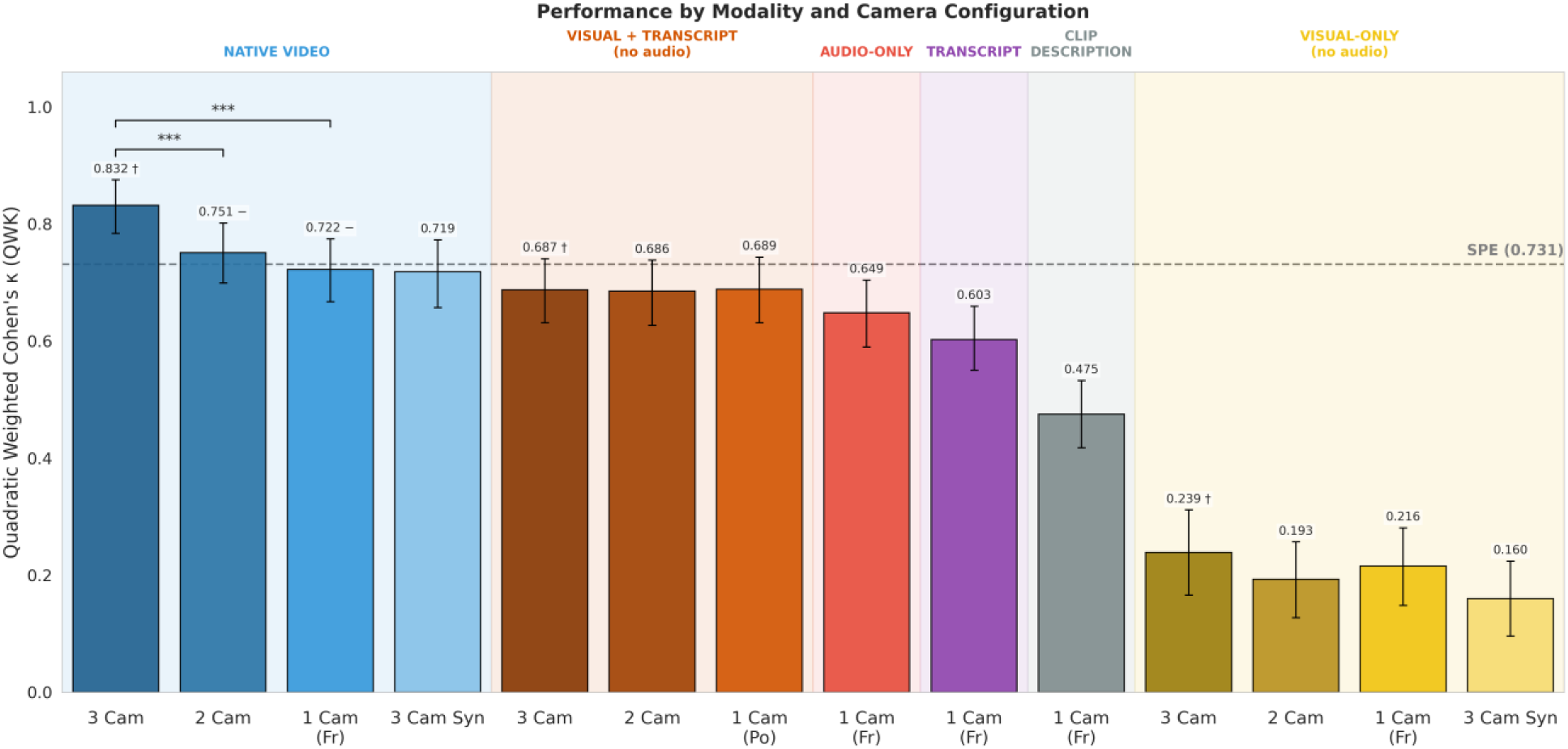
Performance comparison between different configurations via quadratic weighted Cohen’s κ. Brackets denote pairwise differences with p-values < 0.05, relative to the within-modality 3-camera reference (*/**/*** for p < 0.05/0.01/0.001). Dotted line indicates human SPE reference. Results from Gemini 2.5 Pro. Camera configurations: 3 Cam (Fr+Po+Fi), 2 Cam (Fr+Po), 1 Cam (Fr). Abbreviations Fr, Po, Fi, and Syn denote Frontal, Posterior, Fish-eye, and Synthesis.

The performance of multimodal AI varied dramatically according to system input configuration (Figure 2) with an emerging performance hierarchy: Native Video > Audio-Only > Transcript-Only > Visual-Only.

The Gemini 2.5 Pro 3-camera (Fr+Po+Fi) configuration achieved the highest agreement (κ=0.832), significantly outperforming 2-camera (κ=0.752) and 1-camera configurations (κ ≈ 0.72). Differences in audio-only models (κ=0.649) and transcript-only approaches (κ=0.587, p = ns, Table S4) were not statistically significant. While both achieved moderate agreement approaching human SPEs, they lacked the precision of the full video models (p < 0.001, Table 4; Table S4). Visual-only configurations performed the worst, showing low agreement with the Ground Truth (κ ≈ 0.20).

Gemini 2.5 Flash and Qwen-3-omni followed similar trends but with lower overall reliability (Table S3, S7). Notably, Flash’s performance was roughly equivalent across different camera configurations, suggesting it may be less capable of integrating complex multi-view visual data than the Pro model. (Note: A small number of encounters were excluded from Flash analysis due to safety filter refusals; completion rates are detailed in Table S3).

### Station-Specific Performance

When AI model performance is assessed via the macro-F1 score, the hierarchy of modalities remains consistent. However, inspecting performance on individual rubric items reveals configuration-specific differences (Figure 3).

**Figure 3.**
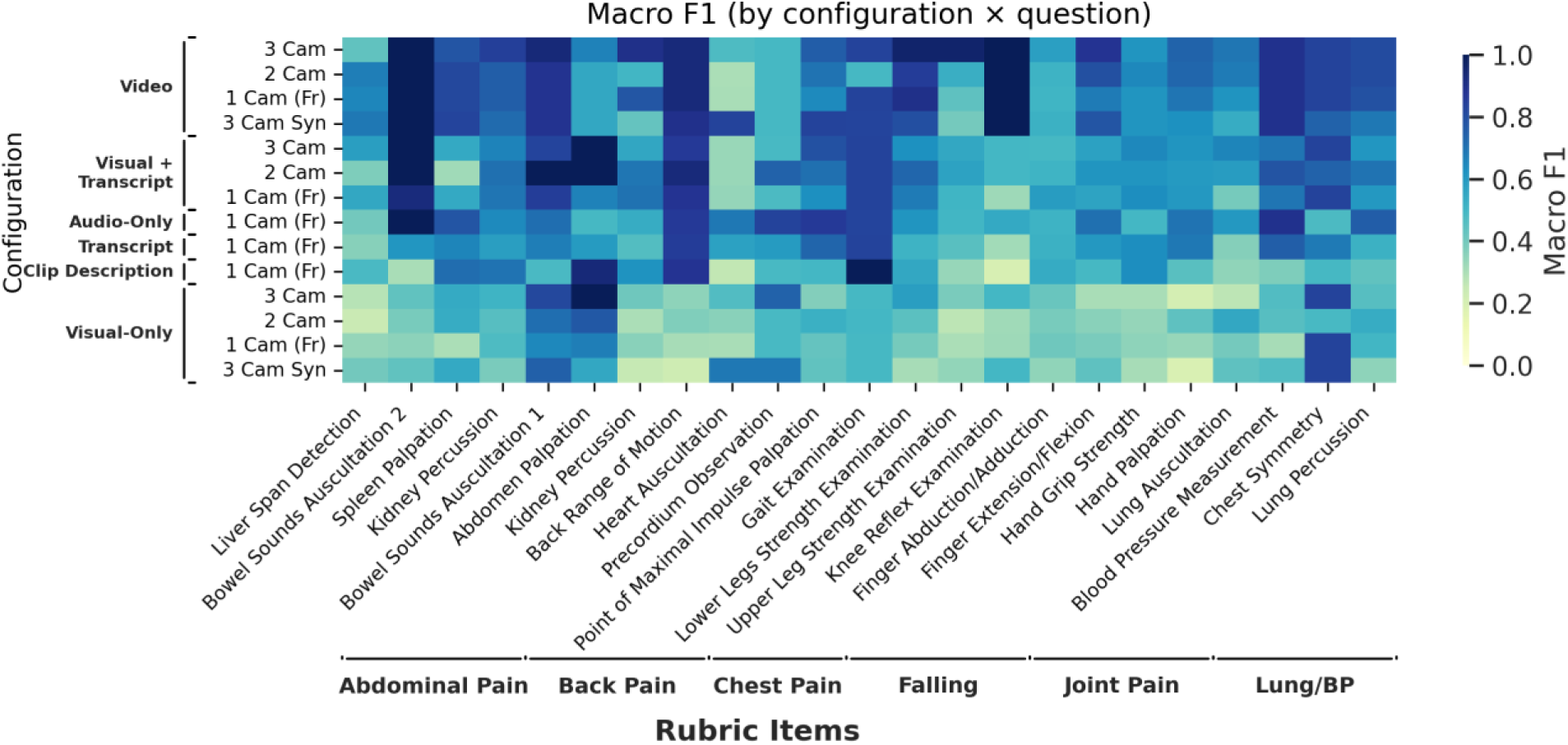
Performance comparison between different configurations and OSCE rubric items via macro F1. Results from Gemini 2.5 Pro.

Native video performed particularly well on tasks emphasizing visual and spatiotemporal action understanding, such as knee reflexes, leg strength, back range of motion, and bowel sound assessment. While native video generally outperformed other setups, Visual+Transcript achieved higher scores on specific items where verbal confirmation aids visual ambiguity, such as abdominal palpation (Visual+Transcript F1 = 1.0 vs. Native Video F1 = 0.66). Models generally underperformed on tasks with subtle cues requiring high-fidelity perception, such as cardiac and pulmonary auscultation (Figure 3, Table S8).

Because multi-camera video provides richer behavioral information but carries computational costs, we compared the 3-camera native video configuration with transcript-only grading. Across the 21 rubric items, native video showed substantially higher performance (mean κ = 0.667 vs 0.395; macro-F1 = 0.777 vs 0.592; agreement = 92.3% vs 79.8%) (Figure 2, Table S4). The largest gains from native video occurred for highly visual tasks such as knee reflexes (Δκ = 1.02), leg strength (Δκ = 0.689), and kidney percussion (Δκ = 0.576).

Transcript-only grading performed best on items with explicit verbalization, including hand-grip strength, abdominal palpation, and gait evaluation, and performed worst on tasks with little or no verbal signposting, including liver span detection and lung auscultation. Transcript-only slightly exceeded native video on a small number of verbally cued items including heart auscultation and precordium observation. These findings indicate that modality performance varies by clinical scenario, with native video favored for visually grounded examinations and transcripts adequate for verbally guided ones.

### The Visual Paradox

Visual-only configurations performed worst across all modalities (κ ≈ 0.20, Table S3). To determine whether this failure stemmed from temporal localization (inability to identify when actions occur) or visual interpretation (inability to understand what actions are), we conducted a targeted re-analysis. From the 1,150 rubric items, we identified the 1,034 (90%) where the native tri-camera model detected an action (score ≠ 0). For these items, we extracted the action timestamps from the tri-camera model’s output and trimmed the muted video to show only the relevant segment for each rubric item.

The standard visual-only configurations (full segmented exam video) achieved 79.5% raw accuracy but poor discrimination (κ = 0.113), primarily by defaulting to the majority class (“Correctly Performed,” 83% of the subset). The targeted visual-only model (trimmed video segments) improved discrimination (κ = 0.150, Wilcoxon signed-rank test, p < 0.001) but raw accuracy dropped to 70.6% (McNemar’s test, p < 0.001; Table S5, S6)—a prevalence paradox.

These results demonstrate that even if the model knows when an action occurs, it cannot reliably determine what occurred from visual information alone. Unlike human observers who can interpret physical examination maneuvers directly from motion, the model appears to require audio (whether through verbal signposting or ambient acoustic cues) to identify and grade examination techniques.

### Inter-rater Reliability

The AI–SPE difference heatmap (Figure 4) shows that with multi-camera video, AI meets or exceeds standardized-patient evaluator (SPE) macro-F1 on many items—especially leg strength, knee reflex, and percussion. Deficits concentrate in observation-heavy heart exam items, where label imbalance depresses κ despite high raw agreement.

**Figure 4.**
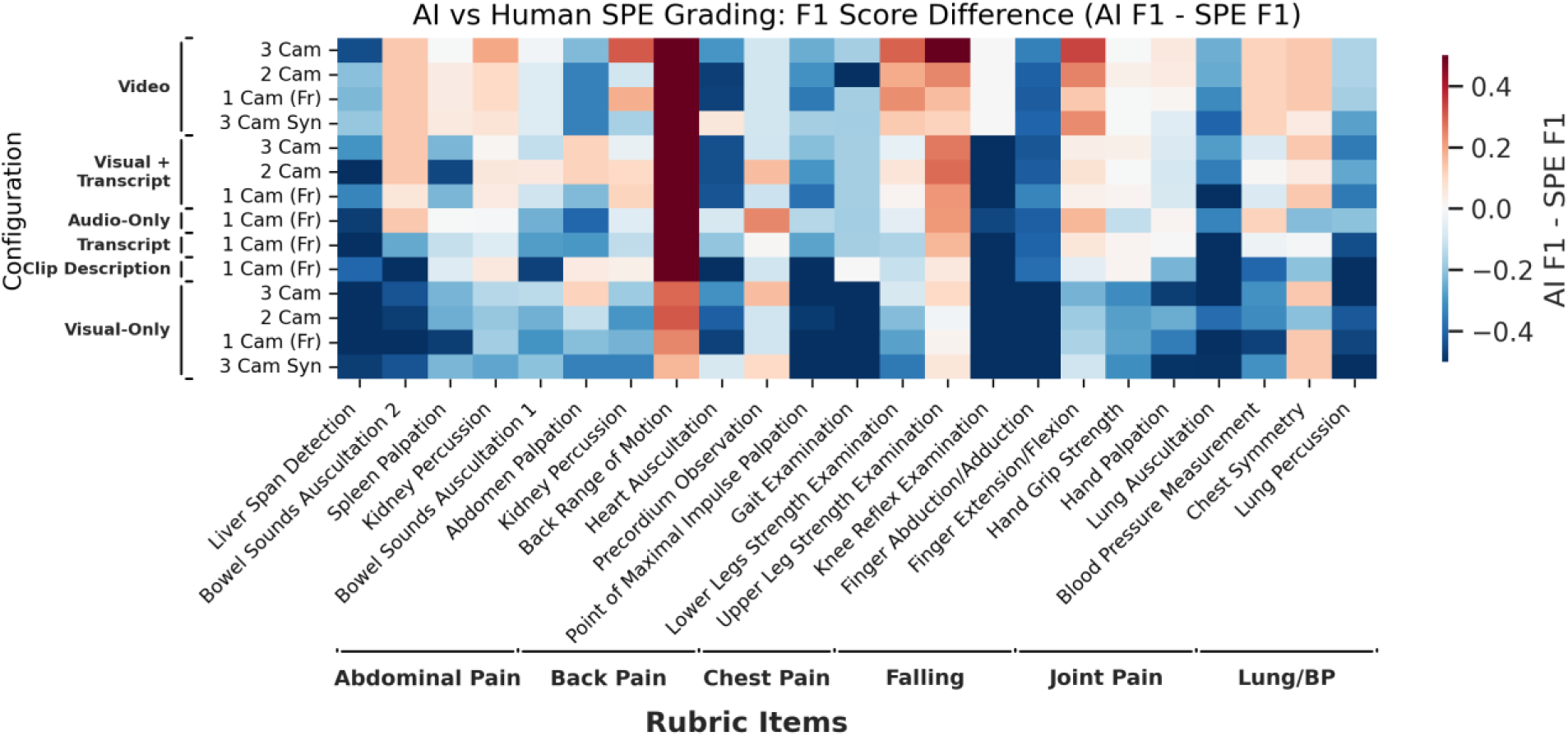
F1 score difference (AI vs. SPE) relative to ground truth across configurations and clinical exams. Red indicates higher AI agreement; blue indicates higher SPE agreement.

Given that human expert graders frequently disagreed with the Ground Truth (κ=0.731), these results underscore the potential of AI-assisted scoring to enhance reliability.

## Discussion

### Principal Findings

In this study of automated OSCE grading focused on physical-examination skill, three elements proved decisive: (i) giving the model native access to synchronized audio and visual streams, (ii) providing multiple coordinated views, and (iii) the consistency of these advantages across stations, with configurations that performed best overall typically performing best for individual maneuvers as well. Together, these ingredients produced the most reliable performance; audio-only was competitive but consistently below full video, transcript-only lagged native audio, and visual-only was least reliable. These patterns are held across stations and rubric items and are consistent with the notion that what is said and what is done each carry distinct, complementary evidence about examination quality.

### Why Native Multimodal Video Works

Direct multimodal processing avoids two failure modes of narrative pipelines. First, converting rich signals (hand placement, bilaterality, contact) into text surrogates discards temporal alignment between what is heard and what is seen; subtle actions that determine credit (e.g., percuss-then-palpate order, on-skin auscultation) are harder to verify once timing is lost. Second, intermediate “stories” are vulnerable to hallucinated detail when the source frames do not fully resolve the action. Operating on the synchronized streams preserves the evidence needed to judge both action and sequence, while the waveform anchors verbal signposting to the corresponding movement. The superiority of native video over transcript-only and the strength of audio-only relative to text-only support this explanation.

### Limitations

This study has several limitations. First, the dataset was collected from a single institution, potentially limiting generalizability to other medical schools with different OSCE formats or student demographics. Second, the “Native Video (3-Camera)” configuration, while the most accurate, represents a significant infrastructural investment that may not be feasible for all centers. Third, while we employed cluster bootstrapping to account for the nested data structure (multiple encounters per student), the sample size of 300 encounters across six stations may not fully capture the long-tail variability of student performance errors. Fourth, our system assumes that student verbal signposting aligns with physical actions; adversarial misalignment, where students describe actions they do not perform, was not tested and could potentially deceive audio-dependent models. Fifth, silent physical examinations without verbal cues would challenge the audio-dependent configurations that performed best in this study. Sixth, results may not generalize to other video-based clinical assessments beyond structured OSCE encounters, such as emergency department evaluations where patient interactions are less predictable. Finally, our analysis focused on physical examination skills; future work should explore the integration of these models with communication skills assessment to provide a holistic view of clinical competence.

## Conclusion

Our systematic ablation analysis demonstrates that AI-driven assessment of physical examination skills is not only feasible but can exceed the reliability of ‘status quo’ grading by human SPEs when provided with the appropriate sensory data. Specifically, the combination of synchronized audio-visual inputs and multiple camera angles is critical for accurately capturing the nuance of “doing” rather than just “saying.” While transcript-based and single-camera approaches offer simpler alternatives, they fail to reach the reliability thresholds necessary for high-stakes assessment. By establishing the “Native Video > Audio > Transcript > Visual” performance hierarchy, this work provides an evidence-based roadmap for medical schools to implement automated grading systems that are both accurate and robust, ultimately paving the way for more frequent, objective, and scalable feedback in medical education.

## Data Availability

All data produced in the present study are available upon reasonable request to the authors

## Acknowledgement

We gratefully acknowledge financial support provided by UT Southwestern institutional funds through the Office of the President. We thank the staff at the UT Southwestern Simulation Center for their invaluable assistance with data collection and for providing access to video recordings of clinical examinations.

